# When will the Covid-19 epidemic fade out?

**DOI:** 10.1101/2020.03.27.20045138

**Authors:** Ilaria Renna

## Abstract

A discrete-time deterministic epidemic model is proposed with the aim of reproducing the behaviour observed in the incidence of real infectious diseases. For this purpose, we analyse a SIRS model under the framework of a small world network formulation. Using this model, we make predictions about the peak of the Covid-19 epidemic in Italy. A Gaussian fit is also performed, to make a similar prediction.

## Introduction

Humanity has always been afflicted by the onset of epidemics. Owing to the absence of vaccines, the slow connections between people and isolation between infectious and susceptible were the only remedies to their devastating effects. Over the last two decades, there have been three major epidemics due to human-transmissible viruses, namely Avian, Ebola and Sars, but fortunately the advanced ability of the scientific world has been able to contain their effects.

A dangerous impact of infectious diseases on populations can arise from emergence and spread of novel pathogens in a population, or a sudden change in the epidemiology of an existing pathogen. Today, due to the absence of a vaccine and to a highly globalized society, the Covid-19 epidemic is frightening the world, raising a series of important questions. Among these, the most common among people is: when will the epidemic die down? During spreading, this is a difficult question to answer: in addition to understand the early transmission dynamics of the infection, control measures should also be accounted for, which may significantly affect the trend of infection.

Generally, the transmission dynamics of an infectious disease is described by modelling the population movements among epidemiological compartments, and assuming random-mixing interactions [1]. When the population mixes at random, each individual has a small and equal chance of coming into contact with any other individual [2]. A more realistic approach describes spatially extended populations, such as elements in a network.

At present, we propose an epidemiological model that describes the population as elements in a network, whose nodes represent individuals while links stand for interactions among them [3]. This model includes the fundamental parameters that characterize a disease, namely infection probability, incubation period of pathogen, social structure and so on.

Using this model and the available statistical data, we attempt predictions on the Covid-19 epidemic trend.

In this short report, we limit ourselves to analyse the epidemiological situation in Italy, and compare to the observed data from France.

### Numerical calculations

Each individual of a population is represented by a node of a small world network [4]. The host population is partitioned into categories containing susceptible *S*, infectious *I*, and recovered individuals *R* (SIRS model). Each node is characterized by a counter *τ*_*i*_. A susceptible individual *S*(*τ*_*i*_ = 0) can come into the infected state through contagion by infected ones. An infected individual *I*(*τ*_*i*_ = 1) passes to the refractory state *R* after an infection time *τ*_*I*_, and a refractory individual returns to susceptible state after immunity duration *τ*_*R*)_ [5]. The cycle is completed after these *τ*_*I +*_ *τ*_*R)*_ time steps, when it returns to the susceptible state. The contagion of susceptible elements occurs stochastically at a local level. If a susceptible element *i* has *k*_*i*_ neighbours, of which *k*_*inf*_ are infected, then it will become infected with probability 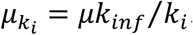. Detail are given in [3].

Figure 1 shows the drawing of the number of infected individuals as a function of time from the beginning of the epidemic until 23 Mars 2020, for a total of 53 days [6]. The active infective individuals are likely underestimated, but this does not change our analysis, as they constitute a subset of the actual ones.

**Figure 1.**
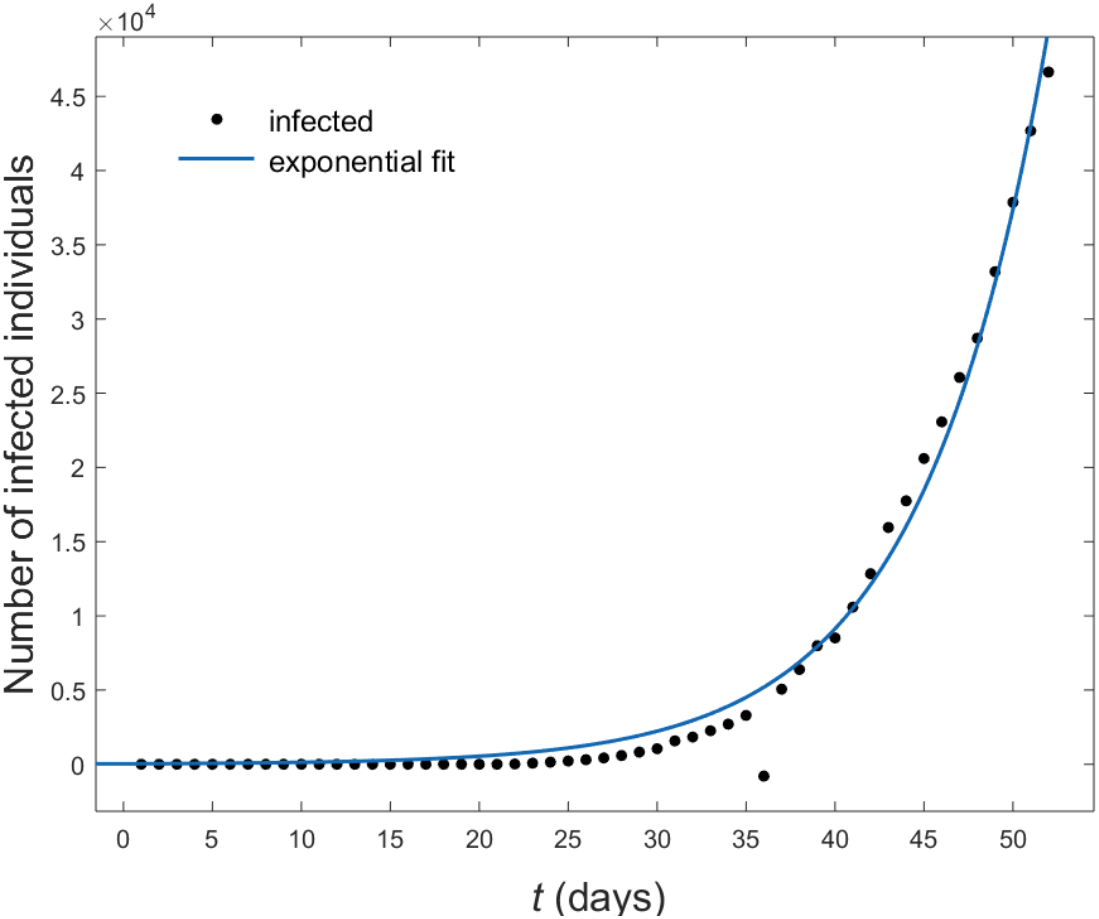
Exponential fit (solid line) of the number of infected individuals as a function of time from the beginning of the epidemic until 23 Mars 2020, in Italy (dots).

As it can be seen from this figure, the trend of the infected is exponential *s (t)* ∝ *e*^*λt*^, with *λ* = 0.13days^-1^. Most likely, this value will decrease, in the coming days, given the very restrictive conditions on the movements of the population enforced by the Italian Government. In Figure 2 we present a realization of our model, wherein the fraction of infected *n*_*inf*_ *(t*_*n*_) is shown, as a function of time, calculated for *N* = 5000 nodes and *p* = 0.1. In the small-world model [4] the parameter *p* controls the transition between a regular lattice (*p* = 0) and the random graph (*p* = 1).

**Figure 2.**
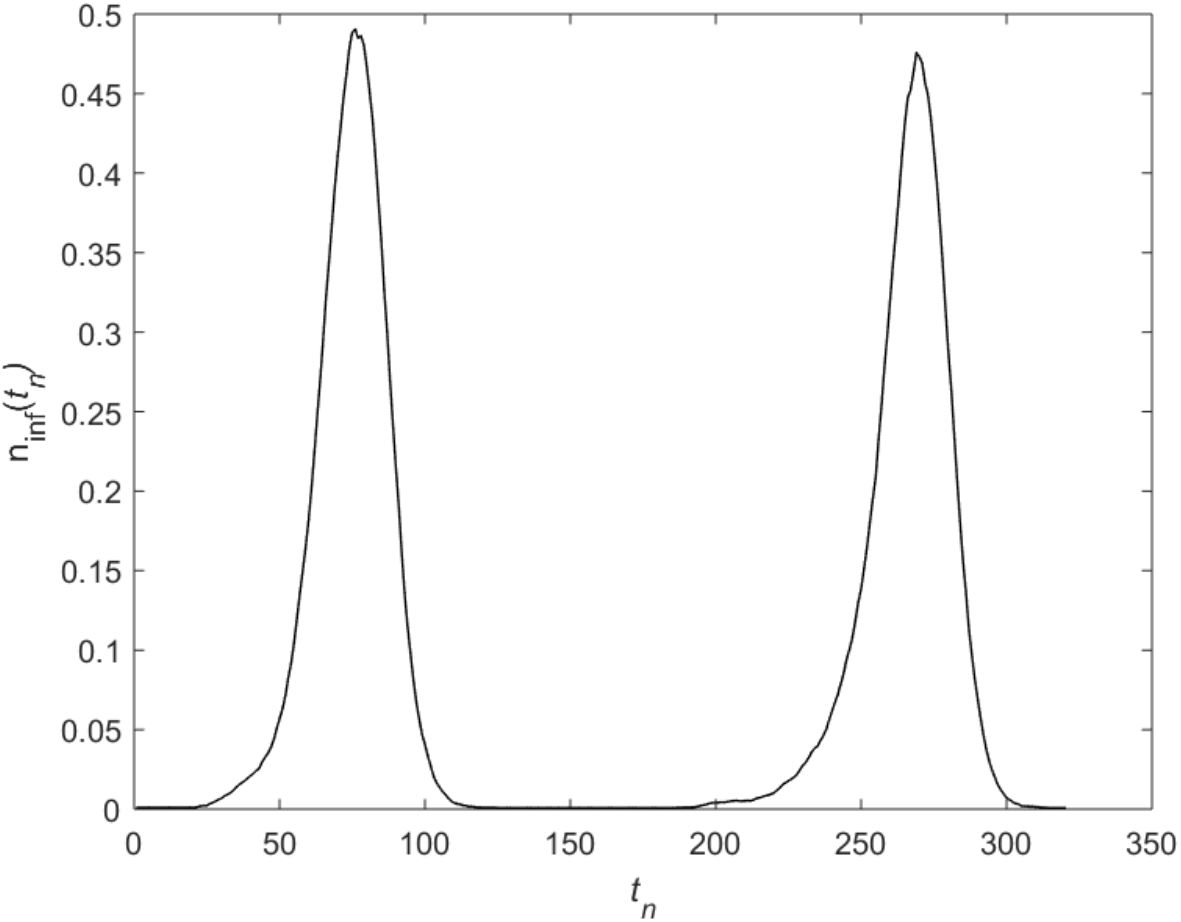
Fraction of infected *n*_*inf*_ *(t*_*n*_) is shown, as a function of time, calculated for *N* = 5000 nodes and *p* = 0.1, *τ*_*I*_ = 14,*τ*_R_ = 150, *μ* = 0.4 and initial fraction of infected *I*_*i*_ = 0.001.

The initial fraction of infected is *I*_*i*_ = 0.001 (that is 5 individuals). The parameter *τ*_*I*_ = 14 was chosen on the basis of median incubation period for COVID-19 estimated in 11.5 days of infection [7]: a larger value was chosen because there can be a delay to symptom appearance resulting from the incubation period. The refractory period was chosen arbitrarily as *τ*_R_ = 150, for calculation purposes only. The network is supposed static, that is no individuals can change their links.

Self-sustained oscillations with a period of nearly 200 days characterize the time series pattern. The system has phase-synchronized oscillations, as it is typical of influenza viruses. As the number of nodes is finite, the infection could end at a fixed time *(t)*, with no further evolution. Therefore, we added sources of infection in the model, i.e. a limited number of infective individuals remain in this state all the time [8].

We are interested in the initial growing part of the figure, that is, the one relating to the first 53 days of epidemic.

In Figure 3 the plot of infected individuals in the first 53 days of disease is compared with the model time series of infected nodes *n*_*inf*_ *(t*_*n*_).

**Figure 3.**
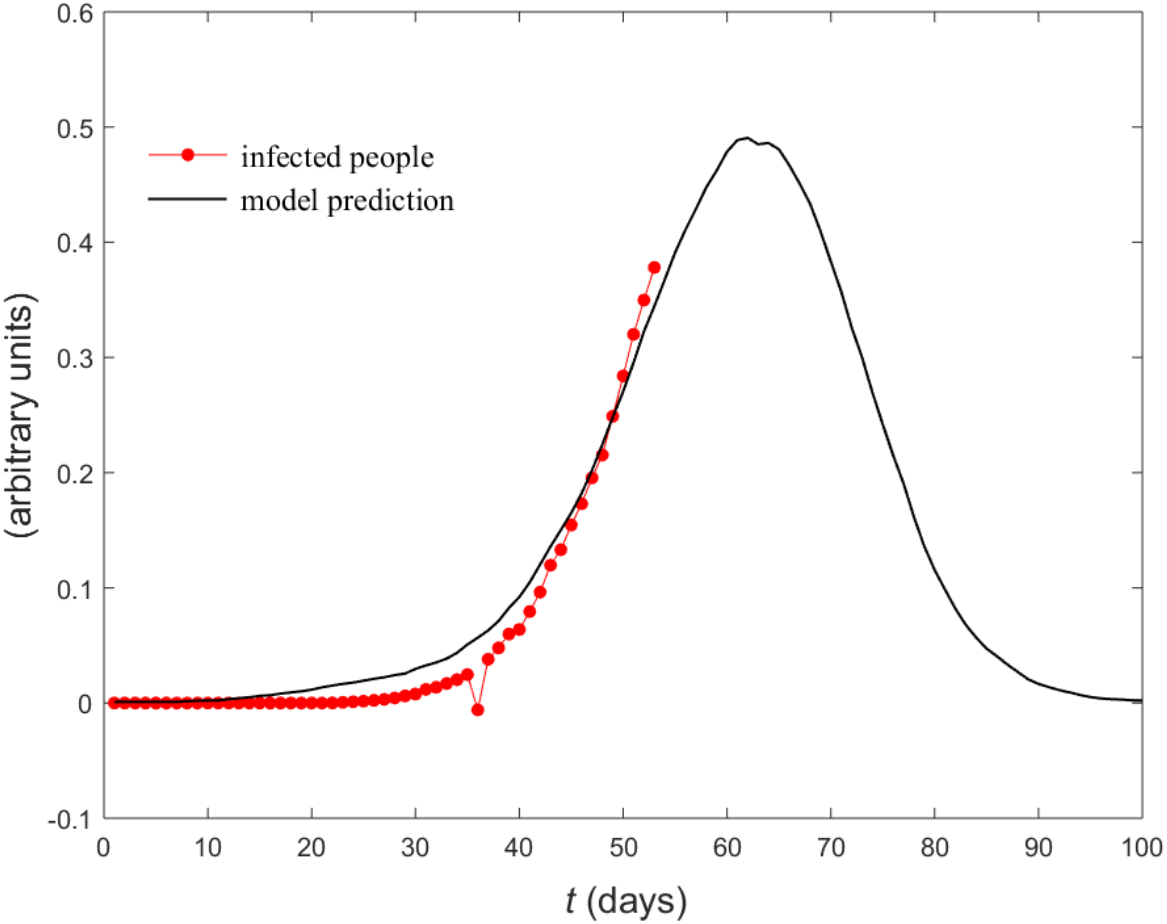
Exponential fit (solid line) of the first 53 time steps of infected nodes fraction (dotted line).

Also in this case, the trend of the infected nodes is exponential 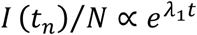, but with *λ*_1_ = 0.11days^-1^. The curve of the infected in the model is less steep than that of the real data and has the first maximum at *t* = 62. Thus, according to the model, the peak of disease will be 62 days after the epidemic outbreak, that is about the 2^nd^ of April 2020. Due to the aforementioned actions taken by the Italian government, it will probably happen before that day.

The shape of each epidemic wave is similar to a Gaussian. Therefore, in Figure 4 a Gaussian curve fit with expected value *μ* = 73 and variance *σ* = 16 is added to the epidemic data. Unlike the previous case, the prediction is that a maximum of 10^*N*^infected will be reached 73 days from the beginning of the epidemic that is about 13 April 2020.

**Figure 4.**
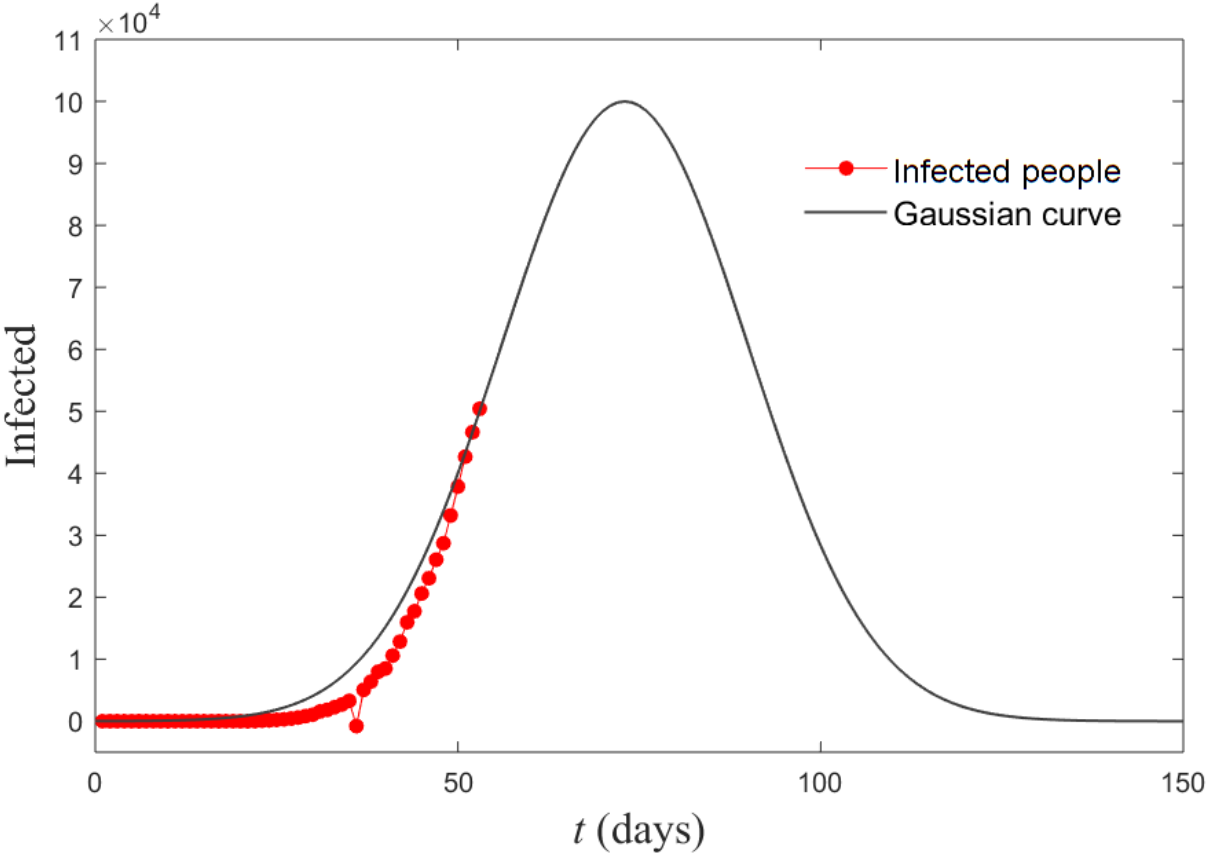
Infected individuals as a function of time (days) until 23 Mars 2020 (dot line), overlapped on a Gaussian curve of the epidemic, with *μ* = 73 and *σ* = 16 (solid line).

We have performed additional calculations allowing the network be rewired. Random rewiring produces larger oscillations, enhancing the maximum number of infected people reached in each period of infectious, whereas adaptive rewiring (as humans tend to respond to the emergence of an epidemic by avoiding contacts with infected individuals) reduces the amplitude of oscillations, accounting for the hoped reduction of infected people [3].

Thus, the effect of Italian Government social distancing orders, aimed at limiting contacts between individuals, should reduce both the value of the maximum in the curve of infected people and the time in which it is reached.

We can therefore say, with an acceptable level of confidence, that the peak in the evolution of the epidemic, with the consequent decrease in infections, should take place between April 2 and April 13, 2020.

The results regarding Italy are also important, because similar epidemic spreads can occur in other countries in Europe. As can be seen in Figure 5, the curve of the infected from Italy and France [9] overlap almost perfectly. Preventive measures taken by other nations could help significantly reduce the spread of the epidemic if taken early.

**Figure 5.**
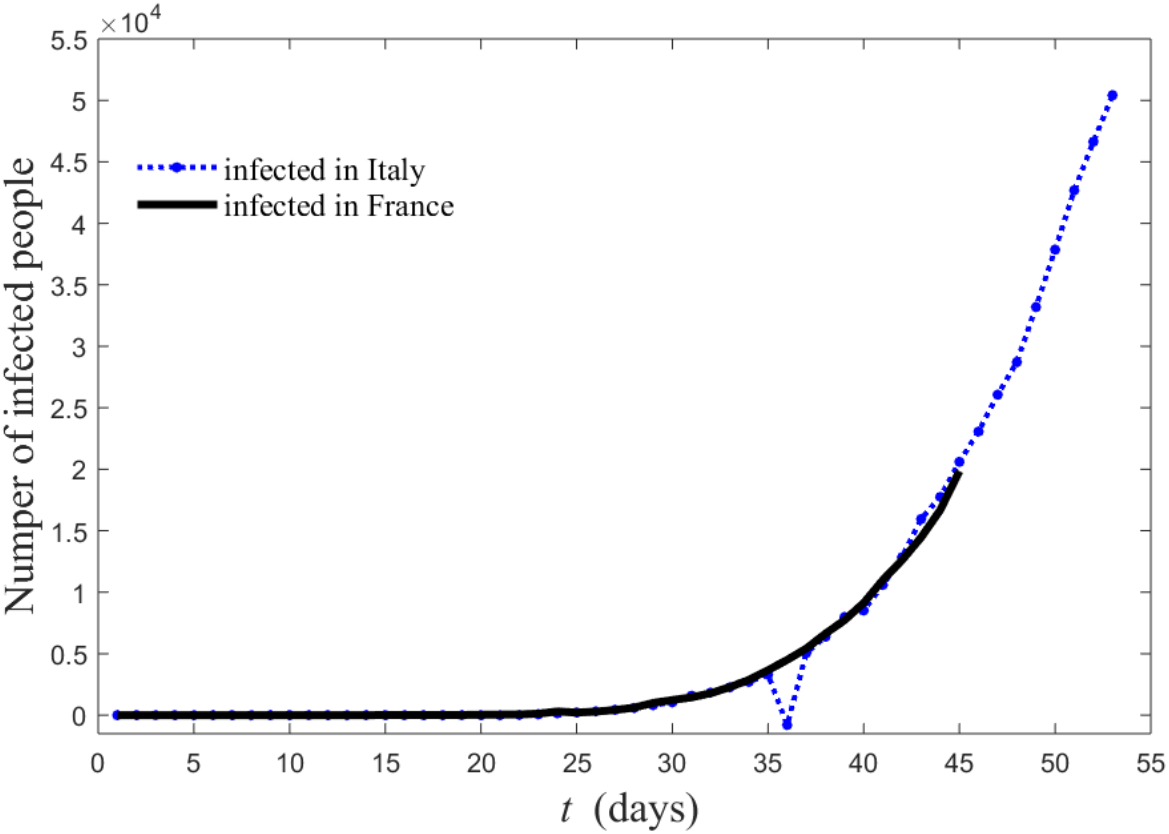
Infected people in Italy (dot line) and in France (solid line). The number of infected in France was shifted back of 10 days.

## Conclusions

We have attempted to describe the pattern of infections of coronavirus disease 2019 (COVID-19) in Italy starting from January 31 to today. Our aim was to make a reasonable prediction on the time the epidemic will reach its peak, with a consequent decrease of disease. We used a SIRS model under the framework of small world network formulation. Incubation period is a fundamental parameter in the epidemic evolution and it can be included directly in the model. We used the model pattern of infected model that fit the real data in the increasing part of initial wave. We estimated the peak as the maximum value in the model curve of infection people. Alternately we used a Gaussian curve, and obtained an additional peak estimation. We estimate that the duration of the COVID-19 disease is around 5 months. This is a rough estimate, as the epidemic spread can be to some extent controlled via contact tracing for suspected cases isolation for confirmed cases.

## Data Availability

All data are available :
https://en.wikipedia.org/wiki/2020_coronavirus_pandemic_in_Italy
https://en.wikipedia.org/wiki/2020_coronavirus_pandemic_in_France

https://en.wikipedia.org/wiki/2020_coronavirus_pandemic_in_Italy

